# Statistical Analysis Plan for Dynamic HIV Choice Prevention delivered with Community Health Workers in Phase A of SEARCH-Sapphire

**DOI:** 10.1101/2023.11.15.23298580

**Authors:** Laura B. Balzer, Elijah R. Kakande, James Ayieko, Jane Kabami, Gabriel Chamie, Moses R. Kamya, Diane V. Havlir, Maya L. Petersen, SEARCH Collaboration

## Abstract

This document provides the statistical analytic plan (SAP) for a cluster randomized trial evaluating the effect of community-based Dynamic Choice HIV Prevention (DCP) intervention, delivered by community health workers in rural Uganda and Kenya (Clinicaltrials.gov: NCT04810650). The SAP was locked prior to unblinding and effect estimation.

## 1. Study Overview

In Phase A of SEARCH-Sapphire (NCT04810650), we are conducting a cluster randomized controlled trial to evaluate the effect of Dynamic Choice HIV Prevention (DCP) delivered by community health workers on biomedical HIV prevention coverage in rural Kenya and Uganda. Details of the trial design and procedures can be found in the corresponding Study Protocol. Analysis plans for qualitative outcomes and cost-effectiveness outcomes are available elsewhere. Power calculations are given in the Appendix.

In brief, we selected 16 villages in rural settings with substantial HIV risk in Kenya and Uganda. These were pair-matched within country on size of village, number of community health workers, and proximity to the highway or a trading center. Then villages were randomized within matched pairs to the DCP intervention or the standard-of-care. Randomization took place at community-based events, where community leaders selected from sealed envelopes containing the trial arm. From May-August 2021, we screened and enrolled 429 participants who were currently or anticipated being at risk of HIV. Follow-up is over 48 weeks.

The DCP intervention includes choice of HIV prevention product (oral pre-exposure prophylaxis [PrEP] or post-exposure prophylaxis [PEP]), choice in HIV testing, choice in service location, and provider training on patient-centered care.

**The primary objective is to evaluate if the DCP intervention improved biomedical HIV prevention coverage, defined as the proportion of the follow-up where the participant self-reported using PrEP or PEP**. Secondary endpoints include biomedical covered time during periods of self-assessed HIV risk (compared between randomized arms) as well as coverage and uptake of the DCP intervention components (within intervention arm only).

## 2. Population and Characteristics

The population of interest is persons aged 15+ years who are residing in study villages, HIV-negative by country-standard rapid testing algorithm, and reporting HIV risk, as assessed via the country-specific Ministry of Health screening tools or self-assessed.

To characterize measurement of this population, we will provide a participant flow diagram (i.e., a CONSORT diagram). Overall and stratified by trial arm and further by sex, we will summarize the baseline characteristics, including sex, age, country, marital status, occupation, HIV risk criteria, alcohol use (any in prior 3 months), mobility (nights away in the past 3 months), pregnancy (women only), circumcision (men only), and any prior use of PrEP or PEP in the past 6 months. We will discretize age into “younger” if aged 15-24 years or “older” if aged 25+ years.

## 3. Endpoint Measurement and Definition

At week-24 and week-48 of follow-up, surveys will be administered to assess HIV risk, possession of PrEP pills, possession of PEP pills, use of PrEP (any doses taken), and use of PEP (any doses taken). The assessment is by month and covers the prior 6 months. We will visualize these data with heatmaps and describe changes in product use over time and by self-reported risk.

The **primary endpoint of biomedical HIV prevention coverage** (a.k.a., biomedical covered time) is the proportion of follow-up where the participant reports taking PrEP or PEP. Thereby, this endpoint has a minimum of 0% (no use) and a maximum of 100% (full coverage). Persons contribute follow-up time when they respond to a survey.

Persons who fail to complete both week-24 and week-48 surveys are missing in the primary analysis. Persons with incident HIV infection are assumed not to be covered during the period prior to seroconversion.

Using these data, we will also define the following **secondary endpoints**:

- Biomedical covered time at-risk, where follow-up is restricted to months of self-reported risk
- Possession covered time, defined as the proportion of follow-up where the participant reports having or receiving PrEP or PEP pills
- Possession covered time at-risk
- Use-to-possession ratio, defined as the proportion of follow-up with PrEP or PEP pills where the participant reports taking them
- Use-to-possession ratio when at-risk

## 4. Evaluation of the SEARCH DCP Intervention Effect

We will assess the Dynamic Choice Prevention intervention effect with targeted minimum loss-based estimation (TMLE), which improves precision and power by adaptively adjusting for baseline outcome predictors.^1–5^ Here, we will use **TMLE with Adaptive Pre-specification** to flexibly control for baseline covariates, while maintaining Type-I error control and accounting for clustering.^6–8^ Using leave-one-village-out cross-validation, we will chose the optimal approach for estimating the outcome regression (i.e., the expected outcome given the randomization arm and adjustment covariates) and the known propensity score (i.e., the conditional probability of being randomized to the intervention given the adjustment covariates). Specifically, we will select the combination of estimators (adjustment variables + approach) that minimizes the cross-validated variance estimate, which again accounts for clustering.

Our pre-specified, candidate adjustment variables consist of sex, age, country, use of PrEP/PEP in the 6 months prior to enrollment, and nothing (i.e., unadjusted). Our pre-specified, candidate learners consist of generalized linear models (GLMs) adjusting for a single variable (beyond the intervention indicator), stepwise regression, multivariate adaptive regression splines (MARS), and the arm-specific mean outcome.

Primary effect estimates will be for the study sample and on the **difference scale**: 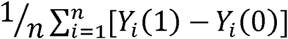, where *n* denotes the total participants, *Y* (*1*) denotes the counterfactual outcome for participant *i* under the intervention and *Y* (*0*) denotes the counterfactual outcome for participant *i* under the control.^9–11^ In other words, we will estimate an individual-level effect (i.e., weight the *n* individuals equally); secondary analyses will estimate a cluster-level effect (i.e., weight the *J* villages equally regardless of their size).^12–14^ Secondary comparisons will be on the ratio scale. All analyses will break the matches used for randomization.^15^

We will test the **null hypothesis** that the intervention did not change biomedical covered time with a two-sided test at the 5% significance level. We will also report point estimates and 95% confidence intervals for each effect measure and the arm-specific outcomes. Standard error estimation will be based on the estimated influence curve, appropriately aggregated to the cluster-level.^14,16,17^ Statistical inference will follow from the Central Limit Theorem,^1^ and will use the Student’s *t-*distribution with (*J - 2*) as a finite sample approximation to the standard normal distribution.^15^

### Secondary analyses

To assess the robustness of these findings, we will repeat these analyses using the unadjusted effect estimator. We will also repeat these analyses using TMLE to adjust for missing endpoints.

### Subgroup analyses

We will repeat these analyses within strata defined by country, sex, age group, alcohol use, and use of PrEP/PEP in the 6 months before enrollment. To further understand effect heterogeneity, we may conduct variable importance measures (i.e., unadjusted and adjusted predictor analyses) with TMLE.

### Secondary endpoints compared by arm

We will implement analogous analyses to evaluate the intervention effect on biomedical covered time at-risk, possession covered time (overall and at-risk), and use-to-possession ratio (overall and at-risk).

### Validation of self-report

Since the primary and key secondary study endpoints rely on self-report, we will objectively measure adherence using drug levels in small hair samples collected among participants reporting any PrEP or PEP doses taken in the past 30 days. Overall and by arm, we will report the number and proportion of these participants with detectable tenofovir levels (>0.002 ng/mg) in their hair at week-24. Using a two-sample test, we will formally test the null hypothesis of equal proportions between arms. We may repeat these analyses at week-48.

### Additional descriptive analyses

Overall and by arm, we will report the number and proportion of participants who withdrew, died, or seroconverted. We will provide seroconversion narratives and may test the null hypothesis of the HIV incidence rate is the same between arms through Poisson regression with person-years-at-risk as offset.

## 5. Intervention Implementation

Within the intervention arm, we will describe coverage and uptake of the DCP intervention over follow-up:

- Visit coverage: number and proportion who attended study visits
- Choice of HIV prevention product: number and proportion who selected PrEP, PEP, condoms, or nothing
- Choice of HIV testing: number and proportion who selected a self-test or rapid HIV test
- Choice of service location: number and proportion who selected to have visits at clinic or at an out-of-clinic location (e.g., home)

All metrics will exclude persons who died, withdrew, or seroconverted by that week of follow-up. At week-48, we will additionally exclude persons who did not reconsent to the extension study (see NCT05549726 for details).

We will also characterize ever use of DCP intervention components over follow-up. We may also report on reasons for product changes, barriers to care, plans to address those barriers, and utilization of the phone hotline. We will report these metrics overall and within key subgroups, such as sex.

## Data Availability

A complete de-identified patient dataset sufficient to reproduce the study findings will be made available approximately one year after completion of the ongoing trial (NCT04810650), following approval of a concept sheet summarizing the analyses to be performed. Further inquiries can be directed to the SEARCH Scientific Committee at.

### Appendix: Power calculations

Sample size and power calculations were based on standard formulas for cluster randomized trials with a continuous outcome.^15^ We expect these calculations to be conservative, because of the precision gained through covariate adjustment during the analysis.

We anticipate an average (harmonic mean) of 20 participants per village. Assuming 10% coverage under the standard-of-care, a standard deviation of 0.35, and coefficient of variation of *k*_*m*_*=*0.25, we anticipate 80% power to detect at least a 15% absolute increase in prevention coverage with *J*=16 clusters (*J*=8 clusters/arm). Even with 20% fewer participants (from 20 to 16 participants/cluster), these calculations suggest we would be well-powered to detect at least a 16.5% absolute increase in prevention coverage.

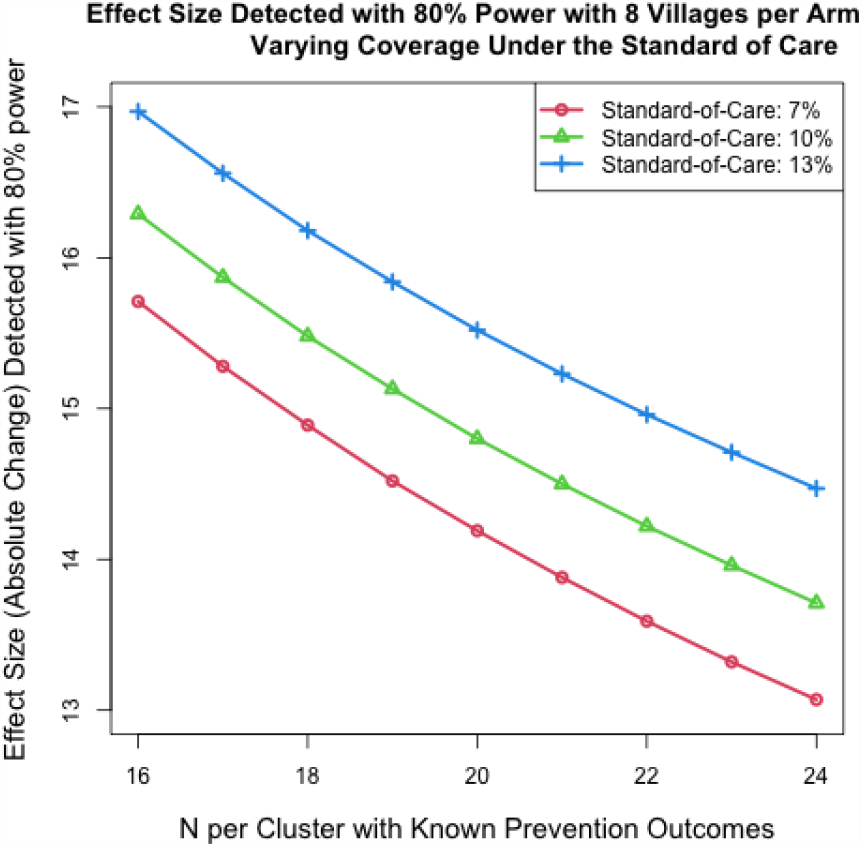

